# Comparative epidemic expansion of SARS-CoV-2 variants Delta and Omicron in Amazonas, a Brazilian setting with high levels of hybrid immunity

**DOI:** 10.1101/2022.09.21.22280193

**Authors:** Ighor Arantes, Gonzalo Bello, Valdinete Nascimento, Victor Souza, Arlesson da Silva, Dejanane Silva, Fernanda Nascimento, Matilde Mejía, Maria Júlia Brandão, Luciana Gonçalves, George Silva, Cristiano Fernandes da Costa, Ligia Abdalla, João Hugo Santos, Tatyana Costa Amorim Ramos, Chayada Piantham, Kimihito Ito, Marilda Mendonça Siqueira, Paola Cristina Resende, Gabriel Luz Wallau, Edson Delatorre, Tiago Gräf, Felipe Naveca, the Fiocruz COVID-19 Genomic Surveillance Network

## Abstract

The SARS-CoV-2 variants of concern (VOCs) Delta and Omicron spread globally during mid and late 2021, respectively, with variable impact according to the immune population landscape. In this study, we compare the dissemination dynamics of these VOCs in the Amazonas state, one of Brazil’s most heavily affected regions. We sequenced the virus genome from 4,128 patients collected in Amazonas between July 1st, 2021 and January 31st, 2022 and investigated the lineage replacement dynamics using a phylodynamic approach. The VOCs Delta and Omicron displayed similar patterns of phylogeographic spread but significantly different epidemic dynamics. The Delta and Omicron epidemics were fueled by multiple introduction events, followed by the successful establishment of a few local transmission lineages of considerable size that mainly arose in the Capital, Manaus. The VOC Omicron spread and became dominant much faster than the VOC Delta. We estimate that under the same epidemiological conditions, the average Re of Omicron was ∼3.3 times higher than that of Delta and the average Re of the Delta was ∼1.3 times higher than that of Gamma. Furthermore, the gradual replacement of Gamma by Delta occurred without an upsurge of COVID-19 cases, while the rise of Omicron fueled a sharp increase in SARS-CoV-2 infection. The Omicron wave displayed a shorter duration and a clear decoupling between the number of SARS-CoV-2 cases and deaths compared with previous (B.1.* and Gamma) waves in the Amazonas state. These findings suggest that the high level of hybrid immunity (infection plus vaccination) acquired by the Amazonian population by mid-2021 was able to limit the spread of the VOC Delta and was also probably crucial to curb the number of severe cases, although not the number of VOC Omicron new infections.

## INTRODUCTION

The SARS-CoV-2 variants of concern (VOC) Delta and Omicron (BA.1) have spread worldwide and reached dominance across geographic regions with different levels of population immunity (Chen et al., 2022; Telenti et al., 2022). A previous study that models the impact of SARS-CoV-2 variants across different scenarios predicted that the population-level impact of new viral variants might vary according to their phenotypes in immunized populations. Viral variants that mainly display enhanced intrinsic transmissibility will lead to small numbers of reinfections/breakthrough infections, which may limit the epidemic’s size. In contrast, viral variants that combine increased transmissibility and immune escape features will produce significant numbers of reinfections/breakthrough infections. In this scenario, the epidemic size is expected to be markedly increased without adapted pharmaceutical and nonpharmaceutical interventions (NPIs) (Bushman et al., 2021).

The immune population landscape may also shape the impact of new SARS-CoV-2 variants. Previous studies demonstrate that “hybrid immunity”, produced by a combination of infection and SARS-CoV-2 vaccination, improves the potency, breadth, and durability of serum neutralizing activity against SARS-CoV-2 variants, including Delta and Omicron, relative to either infection or vaccination alone (Andreano et al., 2021; Bates et al., 2022; Gazit et al., 2022; Goldberg et al., 2022; Hall et al., 2022; Hammerman et al., 2022; Cerqueira-Silva et al., 2022a; Wang et al., 2021; Wratil et al., 2022; Cele et al., 2021; Planas et al., 2021; Rössler et al., 2022; Walls et al., 2022; Gruell et al., 2022; Nielsen et al., 2022; Zar et al., 2022; Altarawneh et al., 2022a). Thus, populations with high hybrid immunity walls are expected to have a better protection against the impact of new SARS-CoV-2 variants carrying immune escape mutations (Crotty, 2021; Suryawanshi & Ott, 2022). Delta and Omicron variant’s relative growth and population impact have been documented in multiple countries with high levels of either natural (like in South Africa) or vaccine (like in the United Kingdom [UK]) acquired immunity (Elliott et al., 2021; Yang & Shaman, 2022; Viana et al., 2022; Elliott et al., 2022; Eales et al., 2022). However, their epidemic dynamics in locations with high levels of hybrid immunity remained poorly described.

The Brazilian state of Amazonas was one of South America’s most heavily affected regions (Faria et al., 2021; Naveca et al., 2021a), and it was estimated that ⩾70% of the population was already infected by March 2021 (He et al., 2021; Buss et al., 2021). At the same time, vaccination roll-out has progressed in the Amazonas since February 2021 (FVS, 2022). Thus, when Delta and Omicron variants were first detected in the state in July and December 2021, respectively (Naveca et al., 2021b; Naveca et al., 2022b), they encountered a population with high levels of preexisting hybrid immunity. This study aimed to compare the population-level impact and dissemination dynamics of SARS-CoV-2 Delta and Omicron variants spreading in the Amazonas state.

## RESULTS

### Temporal and geographic distribution of SARS-CoV-2 VOCs in Amazonas

In this study, we generated 4,128 SARS-CoV-2 high-quality, whole-genome sequences from individuals living in all 11 Amazonas state regions between July 1st, 2021 and January 31st, 2022, representing 3.2% of all laboratory-confirmed SARS-CoV-2 cases (n = 127,787) in the state in that period **(Fig. 1A)**. In line with the geographic location of the SARS-CoV-2 positive cases, most of the genomes were from the Manaus metropolitan area (78%), which comprises the capital city and other surrounding municipalities, followed by the regions of Manacapuru (6%), Parintins (6%), Tefé (3%), and Tabatinga (2%). In comparison, the remaining regions comprise ≤1% of the sequences each (**Fig. 1B**). Out of the 4,128 SARS-CoV-2 genomes generated, 1,024 (25%) were classified as Gamma (P.1/P.1.*), 965 (23%) as Delta (B.1.617.2/AY.*), 2,135 (52%) as Omicron (B.1.1.529/BA.1.*) and four (<1%) as B.1.621. The temporal pattern revealed a gradual replacement of Gamma by Delta followed by a rapid substitution of Delta by Omicron (**Fig. 1C**). Gamma was the most prevalent (>50%) variant in the Amazonas state until mid-September and was detected for the last time on 16^th^ November 2021. Delta was first detected in the state on 21^st^ July 2021 and took ∼70 days to become the most prevalent (>50%) variant and 100-110 days to become the dominant (>90%) variant. Omicron was first detected in Amazonas on 21^st^ December 2021 and rapidly increased in frequency, taking only ∼2 weeks to become the dominant (>90%) variant. The time interval from the first detection to dominance for Omicron (∼10-20 days) was much shorter than for Gamma (∼40-50 days) and Delta (∼100-110 days) variants in Amazonas (**Fig. 1D**). Delta and Omicron were first detected in Manaus city (the Capital) and only later in the countryside municipalities (**Fig. 1E and 1F**). Since the first detection, Delta took ∼80-90 days and >120 days to become dominant in Manaus and outside Manaus, respectively. In comparison, Omicron took <30 days to become the dominant circulating variant within and outside Manaus.

**Figure 1.**
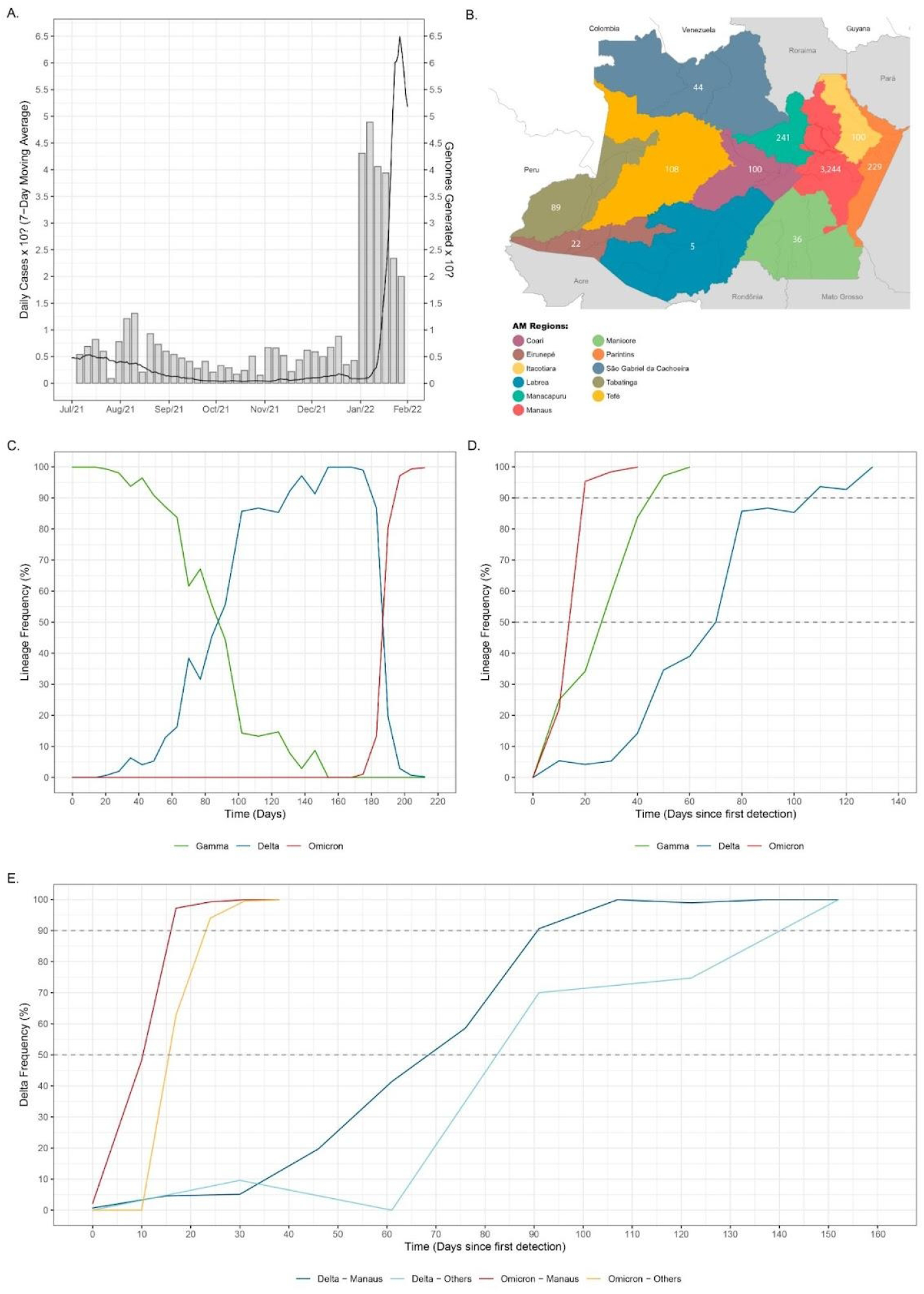
Temporal and geographic distribution of SARS-CoV-2 cases and genomes in Amazonas state. **A)** Rolling mean number of daily SARS-CoV-2 cases (Cota, 2020) in Amazonas between 1st July 2021 and 31st January 2022, along with the number of SARS-CoV-2 genomes sequenced in this study. **B)** Map of Amazonas state showing regions covered by the SARS-CoV-2 genomes generated in this study. Numbers represent genomes generated for each region. **C-D)** Frequency of VOCs Gamma, Delta, and Omicron among SARS-CoV-2 positive samples sequenced in Amazonas since 1st July 2021 (C) or since the first detection of each viral variant **(D). E-F)** Frequency of VOCs Delta (E) and Omicron (F) among SARS-CoV-2 positive samples sequenced within and outside Manaus since the first detection of each viral variant.

### Epidemic dynamics of SARS-CoV-2 VOCs Delta and Omicron

The SARS-CoV-2 epidemic in Amazonas has been characterized by three COVID-19 epidemic waves with exponential growth in cases during the first 24 months **(Fig. 2A)**. The first exponential wave between April and July 2020 was driven by descendants of the B.1 lineage (B.1.195 and B.1.1.28), and the second one between December 2020 and March 2021 by the VOC Gamma. The months following the second wave were characterized by an endemic-like transmission pattern with a roughly constant incidence of ∼100-500 SARS-CoV-2 cases per day. This period was characterized by the gradual replacements of Gamma (P.1) by Gamma descendents lineages (mainly P.1.4 and P.1.6) between April and July 2021 and the subsequent replacement of Gamma descendent lineages by Delta between August and December 2021. This endemic-like period was followed by the third exponential wave that was immediately preceded by the rapid rise of the Omicron variant that fueled a sharp upsurge in the mean number of daily SARS-CoV-2 cases from ∼90 to ∼6,500 during January 2022.

**Figure 2.**
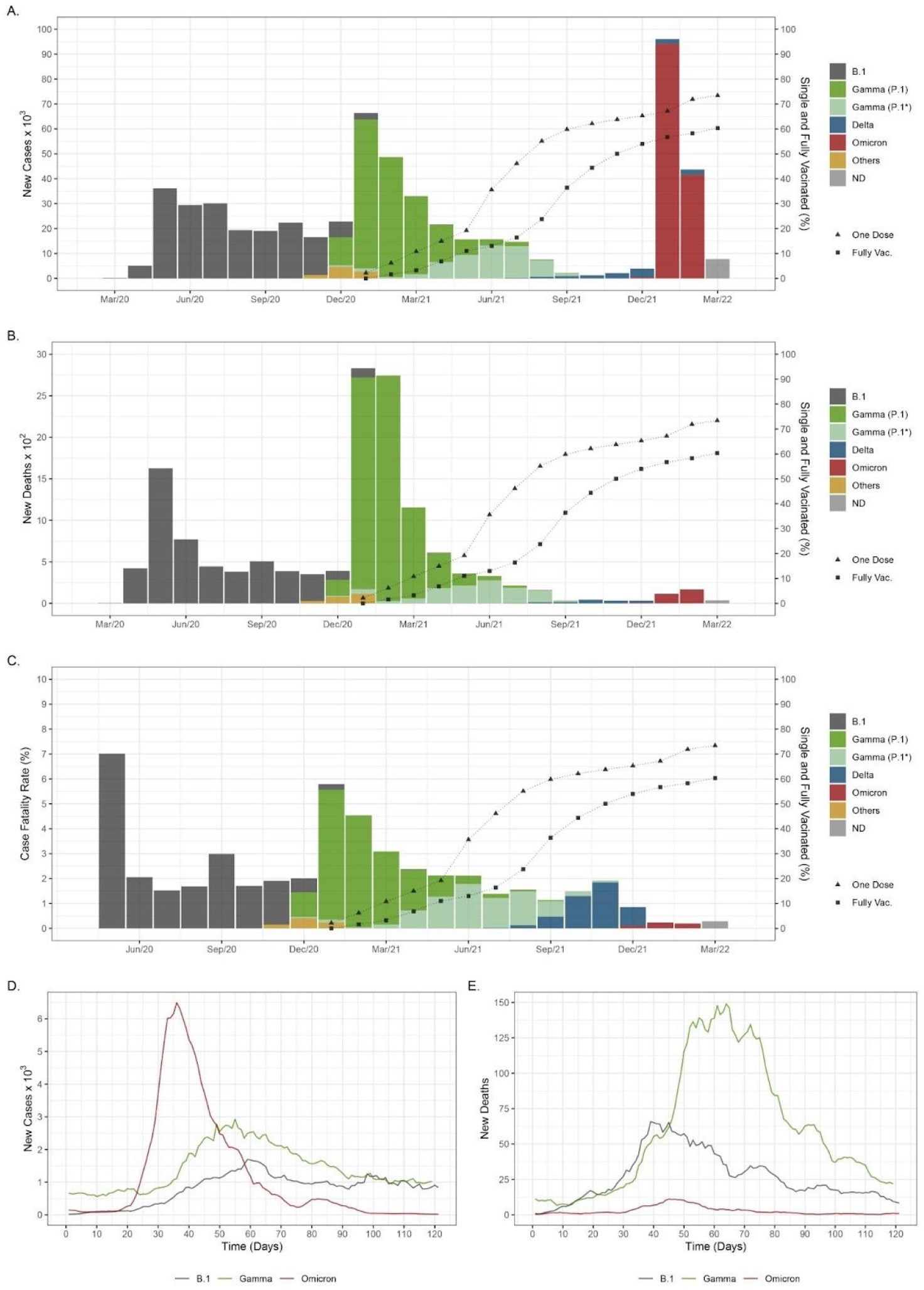
Epidemic trajectories of SARS-CoV-2 variants circulating in Amazonas. **A**. Number of monthly SARS-CoV-2 cases and the percentage of people single and fully vaccinated (Cota, 2020) in Amazonas between March 2020 and March 2022. The color indicates the inferred relative prevalence of viral variants responsible for the infections, as calculated from genomic surveillance data from the EpiCoV database at GISAID. **B**. Number of monthly SARS-CoV-2 deaths and percentage of people single and fully vaccinated in Amazonas between March 2020 and March 2022. **C**. Case-fatality ratio (CFR) is estimated monthly as the ratio between confirmed deaths and cases of SARS-CoV-2 infections and the percentage of people single and fully vaccinated in Amazonas between May 2020 and March 2022. **D-E**. The number of new cases **(D)** and associated deaths **(E)** of B.1, Gamma and Omicron variants in Amazonas since their first detection.

The Omicron wave displayed three significant differences compared with previous waves in the Amazonas state. First, as a result of the rapid rise in infections, the incidence of new daily SARS-CoV-2 cases at the peak of the Omicron wave (6,493) was nearly four-fold and two-fold higher than at the peak of the B.1 (1,696) and Gamma (2,927) waves, respectively **(Fig. 2A and 2D)**. Second, the overall duration of the Omicron wave was much shorter than previous ones. The B.1 and Gamma epidemic waves displayed a mean time of ∼60 days from the onset of case growth to their peaks and another ∼60 days from their peaks to basal levels (<800 daily cases). On the other hand, the peak of the Omicron wave occurred ∼35 days after the first Omicron infection was identified and took another ∼35 days to return to basal levels **(Fig. 2A and 2D)**. Third, the number of deaths during the Omicron wave was much lower than during previous waves **(Fig. 2B and 2E)**, resulting in a substantial reduction in the case-fatality ratio (CFR) **(Fig. 2C)**. The overall CFR estimated during the Omicron wave (0.17) was much lower than that registered during B.1 (3.2) and Gamma (4.1) epidemic waves and was also lower than that observed during the endemic-like period of circulation of Gamma lineages P.1.4/P.1.6 and Delta (1.6-1.7).

### Estimating Delta and Omicron introductions into Amazonas

Most (99%) Delta and Omicron genomes analyzed were grouped in five and nine Pango lineages, respectively (**Table S1**). Of the 965 Delta sequences here analyzed, 496 (52%) were classified as AY.99.2, 147 (15%) as AY.122, 107 (11%) as AY.43, 105 (11%) as AY.9.2, 57 (6%) as AY.101, and 53 (5%) were distributed among other 17 Delta lineages (B.1.617/AY.*). Of the 2,135 Omicron sequences identified, 1,132 (53%) were classified as BA.1, 355 (17%) as BA.1.17.2, 204 (10%) as BA.1.1.15, 115 (5%) as BA.1.9, 82 (4%) as BA.1.14.1, 72 (3%) as BA.1.1, 51 (2%) as BA.1.20, 48 (2%) as BA.1.15, 37 (2%) as BA.14.2 and 39 (2%) were distributed among other eight Omicron lineages (BA.1.*). Lineages AY.99.2 and AY.101 were characterized as being the most prevalent Delta variants of Brazilian origin circulating in the country (Lamarca et al., 2022; Arantes et al., 2022); while lineages BA.1.9 and BA.1.14.* were recognized as BA.1 variants mostly composed by Brazilian sequences (https://cov-lineages.org/lineage_list.html).

To quantify the number of SARS-CoV-2 introductions from abroad into Amazonas, we analyzed sequences of the 14 most prevalent Delta and Omicron lineages combined with their closest worldwide relatives and subjected them to Bayesian phylogeographic analyses (**Fig. S1-S7**). Our analyses detected multiple introductions of Delta and Omicron lineages into Amazonas that peaked in August-September 2021 and the first week of January 2022, respectively **(Fig. 3A and 3B)**. The median estimated number of Omicron (n = 490) introduction events into Amazonas was larger than Delta (n = 120), but greatly varied across lineages ranging from one (95% HPD interval = 1-2) introduction for lineage BA.1.20 to 246 (95% HPD interval = 227-264) introductions for lineage BA.1 **(Fig. 3C and 3D)**. Other Brazilian regions (and particularly the Southeastern region) were pointed as the most important sources of Delta (73%) and Omicron (46%) viruses introduced into Amazonas, followed by neighboring Amazonian South American countries for Delta (18%) and Europe/North America for Omicron (27%/27%) **(Fig. 3E and 3F)**. The region of Manaus, which comprises the capital city and the major national and international airport of the state, received most Delta (84%) and Omicron (91%) introductions from abroad **(Fig. 3E and 3F)**.

**Figure 3.**
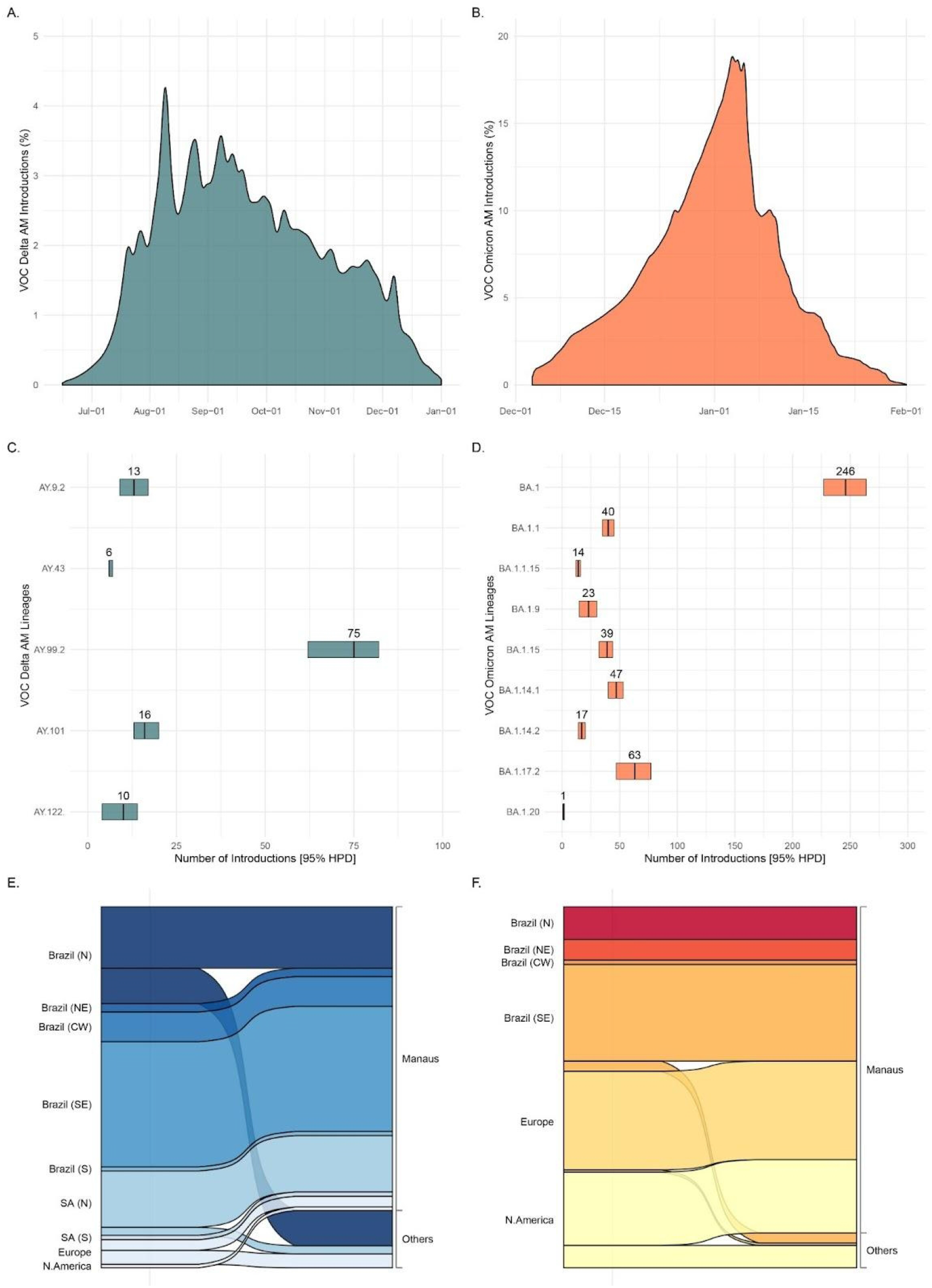
Estimated imports of the most prevalent Delta and Omicron lineages into the Amazonas state. **A-B**. Estimated proportion of introductions of Delta and Omicron into Amazonas state through time, based on the Markov jump history in the Bayesian phylogeographic analysis of the most prevalent Delta and Omicron lineages combined. **C-D**. Median (and 95% credible intervals) estimated number of Delta and Omicron introductions into Amazonas state by lineage. **E-F**. Alluvial plots reflect the total estimated number of Delta and Omicron imports from each source location (left side) into Manaus or other municipalities (right side), inferred from the Bayesian phylogeographic analyses of all main lineages combined. CW: Central-West. N: North. NE: Northeast. S: South. N: North.

### Identification of major Delta and Omicron transmission lineages in Amazonas

Manaus was pointed out as the most relevant source of outgoing Delta (68%) and Omicron (80%) migration events to other Amazonian regions **(Figs. 4A and 4B)**. Most viral introductions in Amazonas, however, showed no evidence of significant onward local transmission and only few of them originated local transmissions lineages of large size defined as highly supported (aLRT > 0.80) monophyletic clades that descend from a most recent common ancestor (MRCA) probably located (PSP [posterior state probability] > 0.90) in Amazonas and that comprises at least 1% of all Delta (n > 10) and Omicron (n > 20) Amazonian sequences. We identified 15 Delta and 17 Omicron Amazonian clades of large size that combined comprise 71% and 47% of all Delta and Omicron sequences from Amazonas here analyzed (**Fig. S1-S7**).. Most Delta (60%) and Omicron (59%) major Amazonian clades belong to lineages AY.99.2 and BA.1 that were mostly (89%) disseminated from other Brazilian regions **(Table S2 and S3)**. In contrast, most Delta and Omicron major clades belonging to other lineages (77%) arose from viruses imported probably from other countries **(Table S2 and S3)**. Most (72%) Amazonian clades of large size probably originated in Manaus and were mostly sampled in this city, a few local clades (6%) arose in Manaus but were more frequently sampled in inner regions, and the remaining clades (22%) arose and were mainly sampled at inner regions of the state **(Figs. 4C and 4D)**. Manaus was the most probable entry point of all major Delta and Omicron lineages introduced from other countries. The lineage AY.99.2AM-IV was the first to become established in Amazonas around late July 2021, but this clade remained restricted to the capital city of Manaus and was last detected in the first week of October 2021. The other major Amazonian Delta lineages only arose after late August 2021 and displayed sustained community transmission until December 2021 to January 2022 **(Fig. 4E)**. All major Amazonian Omicron lineages probably originated between early December 2021 and early January 2022 and displayed sustained transmission until the last week of January analyzed **(Fig. 4F)**. The mean lag in detection since lineage emergence (time of cryptic transmission) was estimated at 13.4 days (SD = 10.7) for Delta lineages and 12.1 days (SD = 9.6) for Omicron lineages **(Figs. 4E and 4F)**.

**Figure 4.**
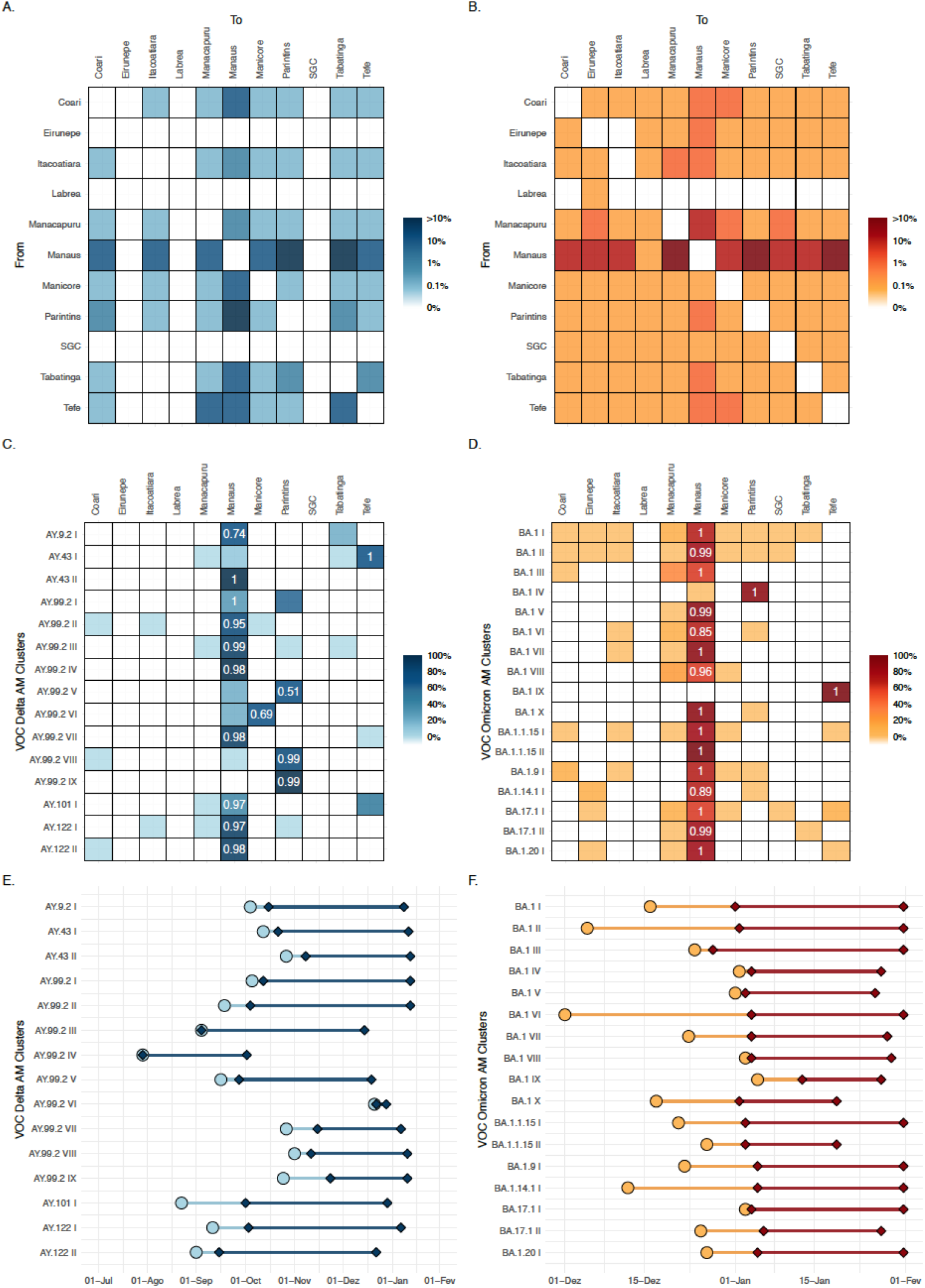
Dissemination of the most prevalent Delta and Omicron lineages within the Amazonas state. **A-B**. Heatmap cells are colored according to the estimated number of Delta and Omicron migrations between Amazonian regions. **C-D**. Heatmap cells are colored according to the proportion of Delta and Omicron sequences from different state regions within each major Amazonian transmission lineage. Numbers indicate the posterior state probability (*PSP*) of the most probable source location. **E-F**. Timeline of major Amazonian Delta and Omicron genomic clusters. Median TMRCA (circles) and dates of first and last sampling times (diamonds) are plotted on the X axis and genomic cluster on the Y axis. Light color lines correspond to detection lag (i.e., the difference between the TMRCA and the date at which a transmission lineage was detected) and dark color lines correspond to the full sampling range of each transmission cluster.

### Effective reproductive number (Re) of SARS-CoV-2 VOCs in Amazonas

The relative Re of VOCs co-circulating in the Amazonas state was first estimated from the observed frequencies of Gamma, Delta, and Omicron from July 2021 to January 2022 using the renewal-equation-based model developed by Ito et al. (Ito et al., 2021). The estimated frequencies suggested that Delta replaced Gamma in September 2021, Omicron replaced Delta at the end of December 2021 **(Fig. 5A)**, and that the replacement from Delta to Omicron resulted in a sudden increase in the average of relative Re of SARS-CoV-2 infections with respect to (w.r.t.) Delta in this region **(Fig. 5B)**. The relative Re of Gamma and Omicron w.r.t. Delta was estimated to be 0.76 (95% CI: 0.74– 0.77) and 3.25 (95% CI: 2.88–3.29), respectively. These results mean that, under the same epidemiological conditions, the average Re of Omicron was 3.25 times higher than that of Delta, the average Re of the Delta was 1.32 times higher than that of Gamma, and the average Re of Omicron was 4.29 times higher than that of Gamma.

**Figure 5.**
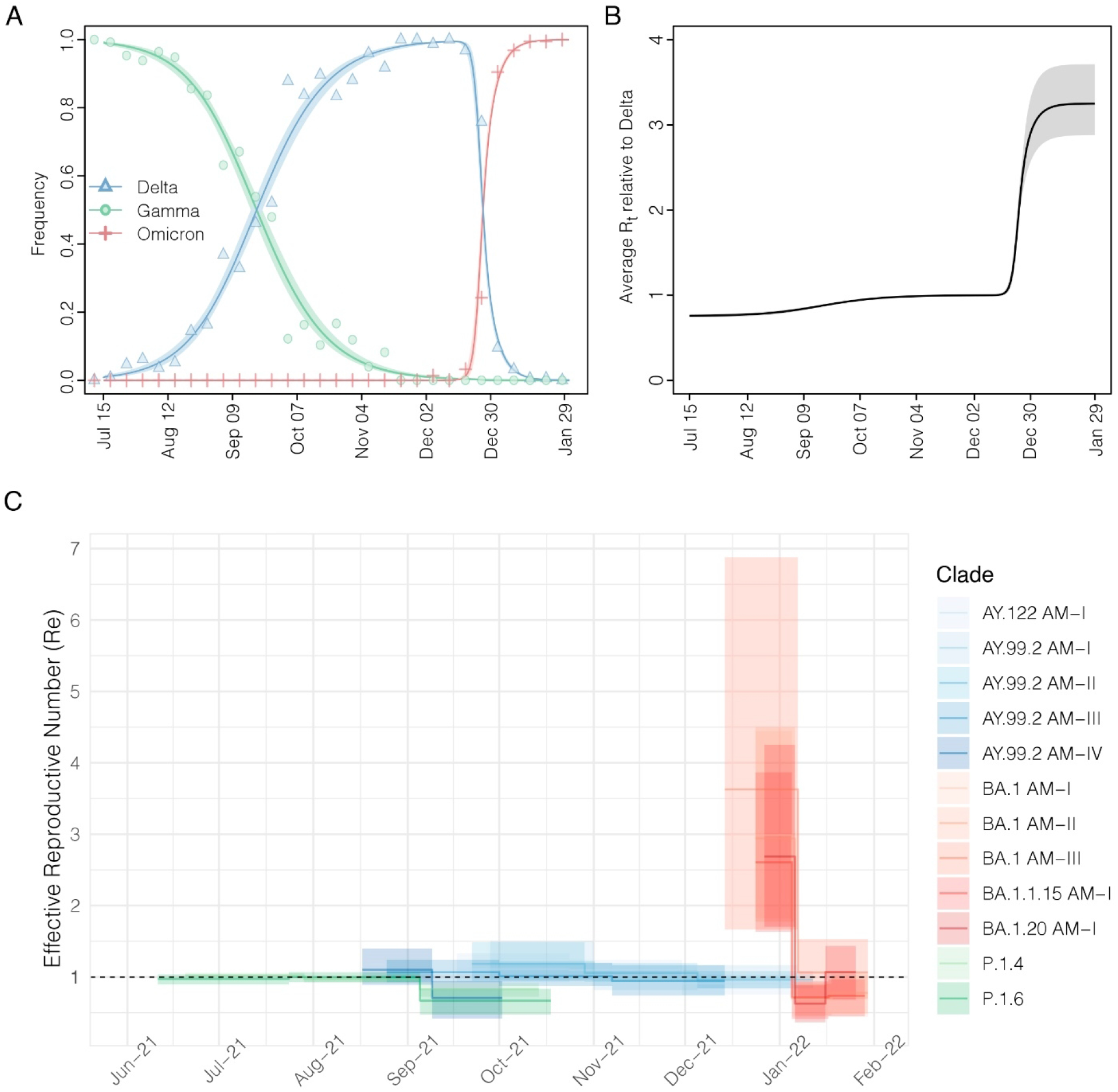
Relative Rt of SARS-CoV-2 VOCs in Amazonas. **A**. Observed and estimated frequencies of SARS-CoV-2 variants in the Amazonas state. Red circles, blue triangles, and orange plus signs represent weekly observed frequencies of Gamma, Delta, and Omicron, respectively. Red, blue, and orange solid lines are estimated frequencies of Gamma, Delta, and Omicron, respectively, and dotted lines represent their 95% confidence intervals. **B**. The estimated average relative Rt of SARS-CoV-2 infections with respect to Delta. Solid lines represent the maximum likelihood estimates, and dotted lines represent their 95% confidence intervals (CI). **C**. Temporal variation in the Re of major SARS-CoV-2 clades circulating in Manaus, estimated using the BDSKY approach. The estimates’ median (solid lines) and 95% HPD intervals (shaded areas) are represented.

We next used the birth-death skyline (BDSKY) model to compare the Re of major SARS-CoV-2 Gamma, Delta, and Omicron lineages circulating in Amazonas. In order to exclude the potential influence of geographical sampling bias, we selected those major Amazonian clusters that originated in Manaus region, and we only included sequences from the capital city. Although the 95% HPD intervals were relatively large, the trajectory of the median Re values was consistent with each lineage’s relative prevalence changes (**Fig. 5C**). Lineages

P.1.* displayed a median Re ∼1 in July and <1 from early August onwards. The Delta lineage AY.99.2_AM-V_ displayed a median Re > 1 in August and then rapidly fell below one in September 2021, preceding subsequent clade extinction. The other Delta lineages (AY.9.2_AM_, AY.99.2_AM-III_, AY.99.2_AM-IV,_ and AY.122_AM_) displayed a median Re > 1 between September and October 2021 and then declined to ≤1 from November onwards. The Omicron lineages displayed a high median Re > 2 during the first weeks after emergence but then rapidly declined to <1 from the second half of January 2022. During the periods of co-circulation of major lineages of different VOCs, the average ratio Re (AY.*)/Re (P.1*) varied between 1.13 and 1.53, and the average ratio Re (BA.1.*)/Re (AY.*) ranged from 2.64 to 3.99.

## DISCUSSION

Our analyses revealed that similar phylogeographic patterns characterized Delta and Omicron epidemics in Amazonas, despite divergent epidemic impacts. The spread of Delta lineages (AY.*) in Amazonas in the second half of 2021 resulted in a gradual replacement of Gamma lineages (P.1.*) without an upsurge of SARS-CoV-2 cases that continued being detected at a roughly steady-state level from July to December 2021. The spread of Omicron lineages (BA.1.*) in Amazonas since late 2021, by contrast, resulted in a rapid replacement of Delta lineages and the exponential growth of SARS-CoV-2 cases. These patterns are entirely consistent with the predicted differential impact of viral variants with enhanced intrinsic transmissibility (like Delta) w.r.t., or variants with combined increased transmissibility and immune escape (like Omicron) spreading in a well-immunized population in the absence of NPIs (Bushman et al., 2021).

Our phylogeographic analyses revealed that Delta and Omicron epidemics derived from multiple introduction events of different lineages. These introductions resulted in a few successful local transmission chains of a considerable size that, combined, comprise 71% and 47% of all Delta and Omicron sequences from Amazonas here analyzed. The number of local large size transmission chains of Delta (n = 15) and Omicron (n = 17) variants here identified, and the mean lag in detection (∼10 days) were roughly comparable. Introductions of both VOCs in Amazonas mostly resulted from domestic viral migration events, followed by migrations from neighboring countries (Peru, Colombia, and Suriname) for Delta lineages and from non-neighboring regions (Europe and North America) for Omicron lineages. The metropolitan region of Manaus acted as the primary entry point and hub of dissemination of both Delta and Omicron variants within Amazonas. Thus, the observed differences between Delta and Omicron epidemics in Amazonas were probably driven by viral characteristics rather than human or stochastic factors.

Two successive VOCs replacements occurred during the study period. Firstly, Gamma lineages (P.1.4 and P.1.6) were substituted by Delta lineages (AY.*), and then Omicron lineages (BA.1.*) replaced Delta sub-lineages in the Amazonas state. Both replacements seem to have been driven by increased virus transmissibility. This result is consistent with previous findings that support the higher infectiousness of Delta compared to Gamma (Du et al., 2022; Y. Liu & Rocklöv, 2022) and of Omicron (BA.1) compared to Delta (Nishiura et al., 2021; Suzuki et al., 2022; Ito et al., 2022; Ito et al., 2022a; Du et al., 2022). Using epidemiological and phylodynamics models, we estimate that the average Re of Omicron was ∼3 times higher than that of Delta. In comparison, the average Re of the Delta was ∼1.3 times higher than that of Gamma sub-lineages in Amazonas. Together, these data explain the much faster lineage replacement of Delta by Omicron than that of Gamma by Delta. Moreover, our results also support that the relative Re under the same conditions predicts the time course of SARS-CoV-2 lineage replacements (Piantham & Ito, 2022). Our results are the first direct comparison between the relative transmissibility of Delta w.r.t. Gamma sub-lineages that harbor the mutations N679K (P.1.4) and P681H (P.1.6). These mutations are particularly interesting because they are located close to the furin cleavage site (FCS) and probably increase viral infectivity (Naveca et al., 2022a). Interestingly, the relative average Re of lineages P.1.4 and P.1.6 w.r.t. P.1 (1.2-1.4) (Naveca et al., 2022a), was similar to the estimated relative transmissibility of Delta w.r.t lineages P.1.4 and P.1.6.

While viral characteristics were the primary determinant of the transmissibility of major Delta and Omicron transmission lineages in Amazonas, other factors have also shaped the dynamics of clade AY.99.2_AM-V_. We observed that the Amazonian clade AY.99.2_AM-V_ was the first large Delta local transmission chain established in Manaus in late July 2021 and displayed a median Re in August that was 1.1 times higher than that of P.1.4 and P.1.6 sub-lineages. During September 2021, however, the Re of clade AY.99.2_AM-V_ fell below one approaching the Re of Gamma sub-lineages, and became extinct in early October 2021. This pattern sharply contrasts with the other large transmission chains that later dominated the Delta epidemic in Manaus and displayed a median Re > 1 during September and October 2021. Our findings revealed that different transmission chains of the same SARS-CoV-2 Delta sub-lineages co-circulating in Manaus displayed variable epidemic expansion and extinction dynamics. Understanding the factors contributing to the early containment and elimination of lineage AY.99.2AM-V in Manaus deserves further investigation.

Although Delta replaced lineages P.1.4/P.1.6 in Amazonas, such variant turnover occurred without a significant upsurge of SARS-CoV-2 cases. This pattern contrasts with the exponential surge of SARS-CoV-2 cases due to the spread of Delta in Europe, North America, Africa, and Asia (Kupferschmidt & Wadman, 2021; Elliott et al., 2021; Dhar et al., 2021; Cherian et al., 2021; Bolze et al., 2022; Yang & Shaman, 2022; Wadman, 2021). However, it coincides with the pattern observed in other Brazilian states and South American countries (Giovanetti et al., 2022; Fonseca et al., 2022). A large fraction of the Amazonian population was probably infected by SARS-CoV-2 (⩾70%) (Buss et al., 2021; He et al., 2021a; Filho et al., 2021; Barber et al., 2022), and another fraction further received at least one dose of the SARS-CoV-2 vaccine (∼50%) previous to the onset of the Delta epidemic (**Fig. 2A**). We suggest that such a high level of hybrid immunity might have prevented the exponential spread of the highly transmissible VOC Delta in Amazonas, and this may explain that the mean Delta’s Re in Amazonas (1.07-1.21) was much lower than the mean Delta’s R0 (5.08) previously estimated (Y. Liu & Rocklöv, 2021). These findings also suggest that the conditional herd immunity reached by South American countries around mid-2021 (Fiori et al., 2022), not before, not only controlled the spread of variants Gamma and Lambda but was also able to protect populations against Delta up-surge. Overall, this supports the notion that the population impact of Delta varies across regions depending on underlying population immunological attributes (Earnest et al., 2022).

Despite several studies pointing to superior immune responses among individuals with hybrid immunity to prevent Omicron infections (Wratil et al., 2022; Cele et al., 2021; Planas et al., 2021; Rössler et al., 2022; Walls et al., 2022; Gruell et al., 2022; Nielsen et al., 2022; Zar et al., 2022; Altarawneh et al., 2022a), the high percentage of people naturally infected and vaccinated (∼65%) (**Fig. 2A**) in Amazonas in December 2021 was not able to prevent the exponential spread of the immune-evasive Omicron variant. This observation is consistent with a recent study showing that even individuals with hybrid immunity had low levels of protection against symptomatic infection by Omicron in Brazil (Cerqueira-Silva et al., 2022b). Furthermore, these results also agree that levels of population immunity that might be sufficient against the Delta variant may not be able to suppress Omicron (Bartha et al., 2021). Of note, the average relative Re of Omicron w.r.t. Delta in Amazonas (∼3) was roughly comparable to that estimated in other settings with different infection and vaccination histories like South Africa, Denmark, the UK, the United States of America, and India (∼2-5) (Nishiura et al., 2021; Suzuki et al., 2022; Ito et al., 2022a; Ito et al., 2022b; Du et al., 2022). The Omicron outbreak in Amazonas also had a similar duration from the onset of case growth to their peaks (∼30 days) than that observed in other populations with different immune landscapes (Washburne et al., 2022; van Dorp et al., 2022). This data supports that immunological-related covariates alone could not explain heterogeneity in Omicron’s fitness across countries (van Dorp et al., 2022).

Currently, it is unclear if the higher transmissibility of Omicron w.r.t to Delta is only mediated by its higher ability to infect individuals with prior immunity to SARS-CoV-2 (Cao et al., 2021; L. Liu et al., 2021; Hoffmann et al., 2022; Eggink et al., 2021; Pulliam et al., 2021; Lyngse et al., 2021; Chaguza et al., 2022) or is also driven by its higher intrinsic transmissibility.

Moderate to high levels of immune evasion, without an additional increase in transmissibility, could explain the observed growth advantage of the Omicron VOC w.r.t Delta in different countries (like South Africa, the UK, Denmark, the USA, and Australia) (Viana et al., 2022; Grabowski et al., 2022). A previous UK study estimated that Omicron is 5–10% less transmissible under a high immune escape scenario than Delta (Barnard et al., 2021). Interestingly, the median estimated Re of BA.1 in Amazonas (Re = 2.53-2.99) and other countries like India (Re = 2.33), South Africa (Re = 2.74-2.79), and the UK (Re = 3.7-4.0) (Y. Liu & Rocklöv, 2022) was lower than the mean estimated Delta’s R0 (∼5) (Y. Liu & Rocklöv, 2021), supporting that the immune evasiveness may primarily drive the rapid spread of Omicron. Previous immunity, however, confers some level of protection against BA.1 infections (Altarawneh et al., 2022a; Altarawneh et al., 2022b; Chemaitelly et al., 2022a; Chemaitelly et al., 2022b; Michlmayr et al., 2022; Andeweg et al., 2022; Lim, 2022), and the Omicron’s R0 should be thus somewhat higher than the estimated Omicron’s Re.

Notably, the median Re of the BA.1 sub-lineages in Amazonas (2.5-3.0) was very similar to the median Re of the B.1.* (Re = 2.1-2.6) and P.1 (Re = 2.6) lineages spreading during the first and second COVID-19 epidemic waves in Amazonas, respectively (Naveca et al., 2021a). Thus, the higher intrinsic transmissibility or immune evasiveness of emergent VOCs surpassed the population immunity levels of the Amazonas population, allowing a new increase in the number of COVID-19 cases, especially during the Omicron wave. Three remarkable differences, however, were observed among waves. The BA.1 wave displayed a higher peak of SARS-CoV-2 cases, shorter duration, and a lower CFR than previous waves. The dissemination of multiple BA.1.* lineages in the absence of mitigation measures may have resulted in a high and sharp peak with a rapid decline, while the curve of previous variants may have been “flattened” and prolonged by control measures implemented at earlier waves. Meanwhile, extensive previous immunity of the Amazonian population likely contributed to decoupling infections and deaths during the BA.1 wave, as observed in other settings (Suzuki et al., 2022; Bager et al., 2022; Davies et al., 2022; Nealon & Cowling, 2022; Arnaout & Arnaout, 2022). Although previous immunity protected against Delta infections, the CRF during the Delta dominant period was higher than during the BA.1 dominant wave, consistent with the notion that Delta is intrinsically more virulent than BA.1 (Bager et al., 2022; Davies et al., 2022; Lewnard et al., 2022; Nyberg et al., 2022; Jassat et al., 2022).

This work has some limitations that should be taken into consideration. First, discrete phylogeographic reconstructions may have been impacted by uneven sampling across geographical locations (Magee & Scotch, 2018; Layan et al., 2022; P. Liu et al., 2022). Second, the inferred number of viral imports into Amazonas could have been affected by sampling coverage and the power to identify transmission clusters from genomic data of variants with extremely rapid global dissemination and low regional diversification (Villabona-Arenas et al., 2020; Gallego-García et al., 2022). We sequenced only a small fraction of infected individuals, and viral imports should thus be interpreted as lower-bound estimates. On the other hand, inferred genomic transmission clusters and some “unclustered” sequences may be part of larger epidemiological transmission clusters, and viral imports may have been thus overestimated. Third, phylogenomic estimates of Re displayed large credibility intervals, making it challenging to obtain precise estimations about relative viral variants’ transmissibility.

In summary, our findings support that the dynamics and population impact of SARS-CoV-2 lineage replacements in Amazonas during the second half of 2021 were driven by virological factors (intrinsic transmissibility and immune-evasiveness of viral strains), combined with levels of prior population immunity. The high level of hybrid immunity (infection plus vaccination) acquired by the Amazonian population by mid-2021 effectively reduced the transmissibility of Delta lineages introduced in the state (Re = 1.1-1.2), resulting in a gradual replacement of Gamma lineages without upsurge of SARS-CoV-2 cases. Such population immunity was not able to prevent the rapid spread of the immune-evasive Omicron BA.1/BA.1.* lineages (Re = 2.0-3.0), which resulted in a fast replacement of Delta lineages and an exponential growth of SARS-CoV-2 cases. However, it was probably crucial to curb the number of deaths during the Omicron wave in Amazonas. Therefore, populations with high levels of hybrid immunity seem to be able to decouple deaths from SARS-CoV-2 infections; but optimization of vaccination strategies will be needed to curtail the transmission of future immune-escape viral variants in those settings.

## METHODS

### SARS-CoV-2 samples and ethical aspects

A total of 4,128 SARS-CoV-2 positive samples with real time RT-PCR cycling threshold (Ct) below 30, collected between 1st July 2021 and 31st January 2022, from residents of 48 out of 62 municipalities from the Amazonas state, were randomly selected for genome sequencing. Nasopharyngeal swabs (NPS) samples collected from suspect COVID-19 cases by sentinel hospitals or health care units were positively tested by real-time RT–PCR using the commercial assay: SARS-CoV2 (E/RP) (Biomanguinhos, https://www.bio.fiocruz.br/index.php/br/produtos/reativos/testes-moleculares/novocoronavirus-sars-cov2/kit-molecular-sars-cov-2-e-rp). The Amazonas State Health Surveillance Foundation - Dra. Rosemary Costa Pinto (FVS-RCP/AM) and the Central Laboratory from the State of Amazonas (LACEN-AM) sent SARS-CoV-2 positive samples for sequencing at FIOCRUZ Amazônia, part of the local health genomics network (REGESAM) and the FIOCRUZ COVID-19 Genomic Surveillance Network. This study was conducted at the request of the SARS-CoV-2 surveillance program of FVS-RCP/AM. It was approved by the Ethics Committee of Amazonas State University (n°. 25430719.6.0000.5016), which waived signed informed consent.

### SARS-CoV-2 amplification, library preparation and genomic sequencing

The viral RNA was subjected to reverse transcription (RT) and PCR amplification using the Illumina COVIDSeq Test (Illumina), including some primers to cover regions with dropout (Naveca et al., 2022a). Normalized pooled amplicons of each sample were used to prepare NGS libraries following manufacturers’ instructions and sequenced in paired-end runs on Illumina MiSeq or NextSeq 1000 platforms at Fiocruz Amazônia.

### SARS-CoV-2 whole-genome consensus sequences and genotyping

Raw data were converted to FASTQ files at Illumina BaseSpace (https://basespace.illumina.com) and consensus sequences were produced with DRAGEN COVID LINEAGE 3.5.5 or 3.5.6. Genomes were evaluated for mutation calling and quality with Nextclade Web 1.13.0 (https://clades.nextstrain.org) (Aksamentov et al., 2021). Sequences with more than 3% of ambiguities “Ns” or any quality flag were reassembled using a customized workflow that employed BBDuk and BBMap tools (v38.84) running on Geneious Prime 2022.0.1. Our consensus sequences had a mean depth coverage higher than 1,800x, excluding duplicate reads. Only those considered as high-quality were considered for further analysis. Whole-genome consensus sequences were classified using the ‘Phylogenetic Assignment of Named Global Outbreak Lineages’ (PANGOLIN) software v3.1.18 (pangolearn 2022-01-20, constellations v0.1.2, scorpio v0.3.16, and pango-designation release v1.2.123) (O’Toole et al., 2021) and later **Selection of SARS-CoV-2 global reference sequences**. To infer introductions for each major SARS-CoV-2 Delta and Omicron lineages circulating in Amazonas, we retrieve all complete SARS-CoV-2 genomes from around the world from the EpiCoV database in GISAID (https://www.gisaid.org/) (Khare et al., 2021) from the respective lineages covering the same time frame than Amazonian sequences (1st July 2021 – 31th January 2022). To select the most closely related reference sequences to the ones in our datasets, a variable number of those with the highest similarity score under a local BLAST (Altschul et al., 1990) search were selected. In all cases, we made sure that reference sequences represented the largest fraction of the final dataset (∼75%). This strategy aimed to keep the total number of sequences within a computationally tractable range (<4,000), but also keep the necessary genomic diversity to discriminate potential dissemination networks.confirmed using phylogenetic analyses.

### Selection of SARS-CoV-2 global reference sequences

To infer introductions for each major SARS-CoV-2 Delta and Omicron lineages circulating in Amazonas, we retrieve all complete SARS-CoV-2 genomes from around the world from the EpiCoV database in GISAID (https://www.gisaid.org/) (Khare et al., 2021) from the respective lineages covering the same time frame than Amazonian sequences (1st July 2021 – 31th January 2022). To select the most closely related reference sequences to the ones in our datasets, a variable number of those with the highest similarity score under a local BLAST (Altschul et al., 1990) search were selected. In all cases, we made sure that reference sequences represented the largest fraction of the final dataset (∼75%). This strategy aimed to keep the total number of sequences within a computationally tractable range (<4,000), but also keep the necessary genomic diversity to discriminate potential dissemination networks.

### Estimating Maximum Likelihood (ML) phylogenetic trees and temporal signal

SARS-CoV-2 Delta and Omicron complete genome sequences from Amazonas were aligned with the global reference datasets of the corresponding lineage using MAFFT v7.467 (Katoh Standley, 2013) and subject to maximum likelihood (ML) phylogenetic analysis using IQ-TREE v2.1.2 (Nguyen et al., 2014) under the general time-reversible (GTR) model of nucleotide substitution with a gamma-distributed rate variation among sites, four rate categories (G4), a proportion of invariable sites (I) and empirical base frequencies (F) nucleotide substitution model, as selected by the ModelFinder application (Kalyaanamoorthy et al., 2017). The approximate likelihood-ratio test assessed the branch support based on the Shimodaira–Hasegawa-like procedure (SH-aLRT) with 1,000 replicates. The temporal signal of the Delta and Omicron assembled datasets was assessed from the ML tree by performing a regression analysis of the root-to-tip divergence against sampling time using TempEst (Rambaut et al., 2016). Preliminary analysis showed minimal temporal signal in most datasets, so all evolutionary analyses (see below) were performed using an informative uniform prior to the molecular clock rate (5-15×10^−4^ substitutions/site/year).

### Estimating time-scaled phylogenetic trees

The time-scaled phylogenetic trees of Delta and Omicron Amazonian transmission clusters were estimated in BEAST v.1.10 (Suchard et al., 2018) under a GTR + F + I + G4 nucleotide substitution model, a strict molecular clock model, and a non-parametric Bayesian skyline (BSKL) model as the coalescent tree prior (Drummond et al., 2005). Due to the large size of Delta and Omicron lineages datasets (n = 372 - 3,445 sequences), time-scaled trees were reconstructed using a modified version of BEAST (https://beast.community/thorney_beast) that makes feasible the analysis of big datasets. The method alleviates most of the computational burden by fixing the tree topology and then re-scales the branch lengths based on clock and coalescent models. The ML phylogenetic trees were inputted in the BEAST xml file as starting, and data trees and analyses were performed as specified above. MCMC was run sufficiently long to ensure convergence (effective sample size> 200) in all parameter estimates as assessed in TRACER v1.7 (Rambaut et al., 2018). The maximum clade credibility (MCC) trees were summarized with TreeAnnotator v1.10 and visualized using FigTree v1.4.4 (https://github.com/rambaut/figtree/releases).

### Discrete Bayesian phylogeographic analyses

A set of 1,000 trees was randomly selected from the posterior distribution of trees resulting from the BEAST analysis of the Delta and Omicron entire lineages datasets. Sampling locations were used as traits in the phylogeographic model, and the ancestral states were reconstructed using a discrete symmetric model with BSSVS (Lemey et al., 2009) on the posterior sampling of trees. SPREAD software (Bielejec et al., 2011) was used to identify the well-supported transition rates based (BF > 3). We complemented this analysis with a Markov jump estimation of the number of location transitions throughout evolutionary history (O’Brien et al., 2009), which was used to explore the directionality of the transitions. Viral introductions were estimated as transitions into Amazonian locations from non-Amazonian locations in the inferred phylogeographic trees, and the date of importation was assumed to be the time of the node inferred to be in Amazonas. Viral migrations within Amazonas were estimated as transitions between Amazonian locations in the inferred phylogeographic trees. Major genomic transmission clusters were defined as highly supported (aLRT > 0.80) monophyletic clades that descend from an MRCA node probably located (PSP > 0.90) in Amazonas and that comprise at least 1% of all Delta (n > 10) and Omicron (n > 20) Amazonian sequences here analyzed. The temporal history of Delta and Omicron imports into Amazonas was summarized as histograms in one-week periods. Supported transitions into Amazonian locations from non-Amazonian ones and transitions among these Amazonian locations were visualized, respectively, in alluvial plots generated with the package “ggaaluvial” (Brunson, 2020), and heatmaps generated with the package “ggplot2” (Wickham, 2016), both available in available in R language (https://www.r-project.org).

### Effective reproductive number (Re) estimations

The relative Re of viral variants in Amazonas was estimated from the observed frequencies of Gamma, Delta, and Omicron from July 2021 to January 2022 using the renewal-equation-based model developed by Ito et al. (Ito et al., 2021). The generation time of Gamma and Delta were assumed to follow a gamma distribution having a mean of 4.7 and a standard deviation of 3.3 (Hart et al., 2022). The generation time of Omicron was assumed to follow a gamma distribution having a mean of 2.8 and a standard deviation of 1.98 (Ito et al., 2022a). We use Delta as the baseline variant to calculate the relative Re of Gamma and Omicron. The relative Re of Gamma and Omicron with respect to (w.r.t.) Delta were estimated by maximizing the multinomial likelihood of the variant frequencies calculated by the model. The 95% confidence intervals of estimated parameters were calculated by the profile likelihood method (Pawitan, 2013). The average of relative generation times and relative Re were calculated by multiplying the frequency and the relative Re of each variant w.r.t. Delta and summing them up (Piantham & Ito, 2022). The temporal Re trajectories of Gamma, Delta, and Omicron variants in Manaus were estimated from genomic data by using the BDSKY model (Stadler et al., 2012) implemented within BEAST 2 v.2.6.2 (Bouckaert et al., 2019). We selected all Amazonian transmission clusters that probably arose in the capital city and comprised at least 40 sequences after removing those sampled outside Manaus. The sampling rate was set to zero for the period before the oldest sample and then estimated from the data afterward. The BDSKY prior settings were as follows: become uninfectious rate (exponential, mean = 36); reproductive number (log-normal, mean = 0.8, s.d. = 0.5); sampling proportion (beta, alpha = 1, beta = 100). Origin parameter was conditioned to root height, and Re was estimated in a piecewise manner over 3-6 time intervals defined from the date of the most recent sample up to the root of the tree. For each transmission cluster tMRCA we applied a uniform prior whose limits were based on the 95%HPD of the estimated origin according to the phylogeographic analysis. MCMC chains were run until all relevant parameters reached ESS >200, as explained above.

## Data Availability

All the SARS-CoV-2 genomes generated and analyzed in this study are available at the EpiCoV database in GISAID (https://www.gisaid.org/) and could be assesed at the following EPI_SET ID: https://epicov.org/epi3/epi_set/220913va?main=true.

## AUTHORS CONTRIBUTIONS

IA, GB, and FGN conceived and designed the study and contributed to data analysis. VN, VS, AS, DS, FN, MM, MJB, LG, and GS contributed to diagnostics and sequencing analysis. CFC, LA, JHS, and TCAR contributed to patient and public health surveillance data. CP and KI contributed to effective reproductive number (Re) estimations. FGN and MMS contributed to laboratory management and obtaining financial support. PCR, GLW, ED, and TG contributed to formal data analysis. IA and GB wrote the first draft, and all authors contributed and approved the final manuscript.

## ACKNOWLEDGMENTS

The authors wish to thank all the health care workers and scientists who have worked hard to deal with this pandemic threat. We also appreciate the support of FIOCRUZ COVID-19 Genomics Surveillance Network members and the Respiratory Viruses Genomic Surveillance Network of the General Laboratory Coordination (CGLab) of the Brazilian Ministry of Health (MoH), Brazilian Central Laboratory States (LACENs), and the Amazonas surveillance teams for the partnership in the viral surveillance in Brazil.

