## Supplementary Material for "Comparative epidemic expansion of SARS-CoV-2 variants Delta and Omicron in Amazonas, a Brazilian setting with high levels of hybrid immunity"

### SUPPLEMENTARY TABLES

**Table S1:** Pango lineages detected in Amazonas state.

| | Lineage | N | N (%) |
|---|---|---|---|
| **Delta** | AY.4 | 1 | <1 |
| | AY.6 | 1 | <1 |
| | AY.9.2 | 105 | 11 |
| | AY.20 | 4 | <1 |
| | AY.34 | 19 | 2 |
| | AY.34.1 | 2 | <1 |
| | AY.34.1.1 | 4 | <1 |
| | AY.34.2 | 1 | <1 |
| | AY.42 | 5 | 1 |
| | AY.43 | 107 | 11 |
| | AY.43.1 | 1 | <1 |
| | AY.59 | 1 | <1 |
| | AY.99.1 | 1 | <1 |
| | AY.99.2 | 496 | 51 |
| | AY.100 | 3 | <1 |
| | AY.101 | 57 | 6 |
| | AY.103 | 2 | <1 |
| | AY.116 | 1 | <1 |
| | AY.119 | 2 | <1 |
| | AY.119.1 | 3 | <1 |
| | AY.122 | 147 | 15 |
| | B.1.617.2 | 2 | <1 |
| | **Total** | **965** | **100** |

| | Lineage | N | N (%) |
|---|---|---|---|
| **Omicron** | BA.1 | 1,132 | 53 |
| | BA.1.1 | 72 | 3 |
| | BA.1.1.10 | 1 | <1 |
| | BA.1.1.14 | 11 | 1 |
| | BA.1.1.15 | 204 | 10 |
| | BA.1.1.18 | 2 | <1 |
| | BA.1.9 | 115 | 5 |
| | BA.1.14.1 | 82 | 4 |
| | BA.1.14.2 | 37 | 2 |
| | BA.1.14 | 19 | 1 |
| | BA.1.15 | 48 | 2 |
| | BA.1.16 | 1 | <1 |
| | BA.1.17 | 4 | <1 |
| | BA.1.17.2 | 355 | 17 |
| | BA.1.18 | 1 | <1 |
| | BA.1.20 | 51 | 2 |
| | **Total** | **2,135** | **100** |

*The table details the absolute and relative frequencies of the Pango lineages to which sequences generated in Amazonas state between July-2021 and March-2022 were assigned. Only sequences in the VOCs Delta and Omicron were listed.*

**Table S2**: Major clades of the VOC Delta detected in Amazonas state.

| Lineage | Cluster | N (%) | Entry Point (PSP) | Source (PSP) |
|---|---|---|---|---|
| AY.9.2 | AY.9.2$_{\text{AM-I}}$ | 88 (84) | Manaus (74) | SA North (91) |
| AY.43 | AY.43$_{\text{AM-I}}$ | 32 (30) | Tefé (100) | EU (99) |
| | AY.43$_{\text{AM-II}}$ | 30 (28) | Manaus (100) | SA North (45) |
| AY.99.2 | AY.99.2$_{\text{AM-I}}$ | 95 (19) | Manaus (100) | BR-SE (92) |
| | AY.99.2$_{\text{AM-II}}$ | 85 (17) | Manaus (95) | BR-SE (96) |
| | AY.99.2$_{\text{AM-III}}$ | 50 (10) | Manaus (99) | BR-CW (100) |
| | AY.99.2$_{\text{AM-IV}}$ | 40 (8) | Manaus (98) | BR-SE (100) |
| | AY.99.2$_{\text{AM-V}}$ | 24 (5) | Parintins (51) | BR-N (70) |
| | AY.99.2$_{\text{AM-VI}}$ | 23 (5) | Manicore (69) | BR-N (97) |
| | AY.99.2$_{\text{AM-VII}}$ | 22 (4) | Manaus (98) | BR-N (99) |
| | AY.99.2$_{\text{AM-VIII}}$ | 19 (4) | Parintins (99) | BR-N (86) |
| | AY.99.2$_{\text{AM-IX}}$ | 16 (3) | Parintins (99) | BR-N (76) |
| AY.101 | AY.101$_{\text{AM-II}}$ | 45 (79) | Manaus (97) | BR-S (99) |
| AY.122 | AY.122$_{\text{AM-I}}$ | 100 (68) | Manaus (97) | SA North (88) |
| | AY.122$_{\text{AM-II}}$ | 17 (12) | Manaus (98) | SA North (62) |

*The table details the major clusters detected in Amazonas state of the VOC Delta, defined by their statistical support (aLRT > 0.80), location of their MRCA (PSP of Amazonas location > 0.80), and dimension (at least 1% of all sequences from the VOC). For each cluster are supplied absolute and relative dimensions, most probable entry point among Amazonas's regions, and the most probable origin of the migration it originated from, the last two accompanied by their PSP values. BR: Brazil, EU: Europe, SA: South America, N: north, S: south, SE: southeast, CW: central-west.**FIGURE***

**Table S3**: Major clades of the VOC Omicron detected in Amazonas state.

| Lineage | Cluster | N (%) | Entry Point (PSP) | Source (PSP) |
|---|---|---|---|---|
| BA.1 | $BA.1_{AM-I}$ | 185 (16) | Manaus (100) | BR-SE (78) |
| | $BA.1_{AM-II}$ | 168 (15) | Manaus (99) | BR-SE (78) |
| | $BA.1_{AM-III}$ | 55 (5) | Manaus (100) | BR-SE (78) |
| | $BA.1_{AM-IV}$ | 49 (4) | Parintins (100) | BR-SE (95) |
| | $BA.1_{AM-V}$ | 38 (3) | Manaus (99) | EU (99) |
| | $BA.1_{AM-VI}$ | 37 (3) | Manaus (85) | EU (100) |
| | $BA.1_{AM-VII}$ | 29 (3) | Manaus (100) | BR-SE (78) |
| | $BA.1_{AM-VIII}$ | 27 (2) | Manaus (96) | BR-SE (81) |
| | $BA.1_{AM-IX}$ | 27 (2) | Tefé (100) | BR-SE (83) |
| | $BA.1_{AM-X}$ | 24 (2) | Manaus (100) | BR-SE (78) |
| BA.1.1.15 | $BA.1.1.15_{AM-I}$ | 165 (81) | Manaus (100) | EU (100) |
| | $BA.1.1.15_{AM-II}$ | 26 (13) | Manaus (100) | EU (100) |
| BA.1.9 | $BA.1.9_{AM-I}$ | 35 (30) | Manaus (100) | BR-SE (70) |
| BA.1.14.1 | $BA.1.14.1_{AM-I}$ | 21 (26) | Manaus (89) | BR-SE (80) |
| BA.17.2 | $BA.17.2_{AM-I}$ | 50 (14) | Manaus (100) | EU (83) |
| | $BA.17.2_{AM-II}$ | 20 (6) | Manaus (99) | EU (83) |
| BA.1.20 | $BA.1.20_{AM-I}$ | 50 (98) | Manaus (100) | BR-SE (97) |

*The table details the major clusters detected in Amazonas state of the VOC Omicron, defined by their statistical support (aLRT > 0.80), location of their MRCA (PSP of Amazonas location > 0.80), and dimension (at least 1% of all sequences from the VOC). For each cluster are supplied absolute and relative dimensions, most probable entry point among Amazonas's regions, and the most probable origin of the migration it originated from, the last two accompanied by their PSP values. BR: Brazil, EU: Europe, N: north, SE: southeast.*

### SUPPLEMENTARY FIGURES:

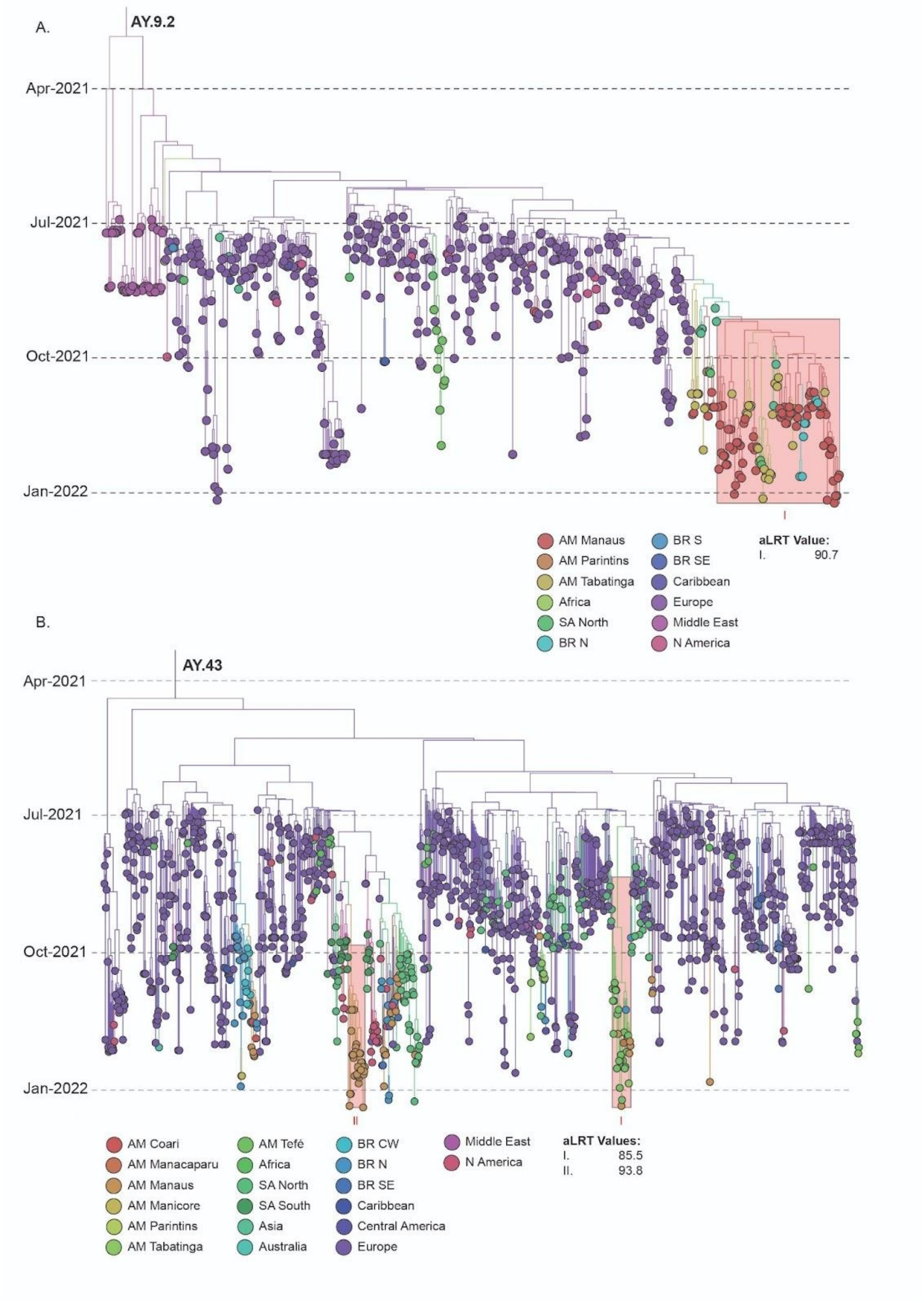


**Figure S1: Spatio Temporal Characterization of lineages AY.9.2 and AY.43 in Amazonas State**. Time-scaled MCC tree of SARS-Cov-2 lineages AY.9.2 ($n_{TOTAL}$ = 616) and AY.43 ($n_{TOTAL}$= 1,339). Branches are colored according to their most probable location, based on the color scheme shown in the legends. Large clusters identified in each lineage are indicated, as are their aLRT support obtained in the previous ML analysis. AM: Amazonas, N: north, S: south, SA: South America, SE: southeast, CW: central-west.

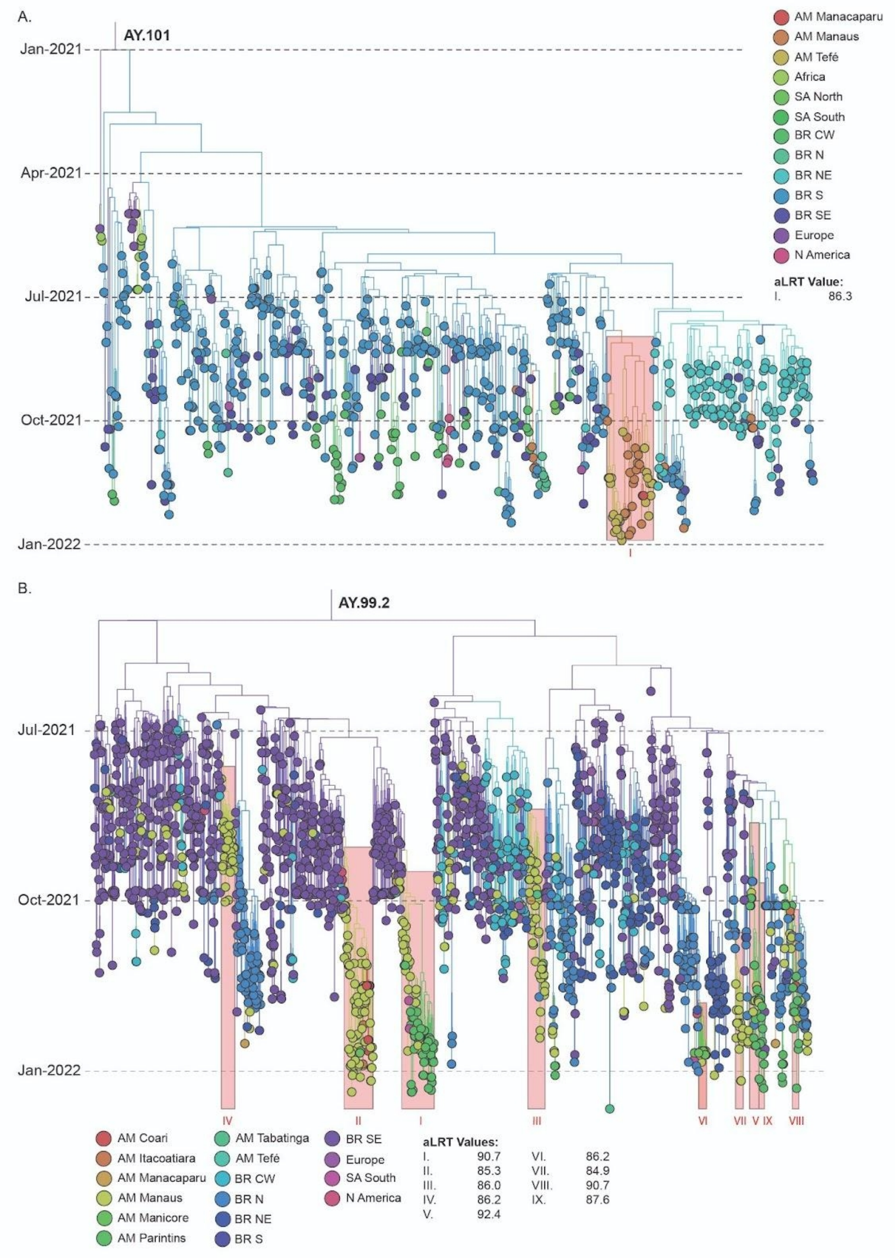


**Figure S2: Spatio Temporal Characterization of lineages AY.101 and AY.99.2 in Amazonas State.** Time-scaled MCC tree of SARS-Cov-2 lineages AY.101 ($n_{TOTAL}$ = 691) and AY.99.2 ($n_{TOTAL}$= 2,102). Branches are colored according to their most probable location, based on the color scheme shown in the legends. Large clusters identified in each lineage are indicated, as are their aLRT support obtained in the previous ML analysis. AM: Amazonas, N: north, S: south, , SA: South America, SE: southeast, CW: central-west.

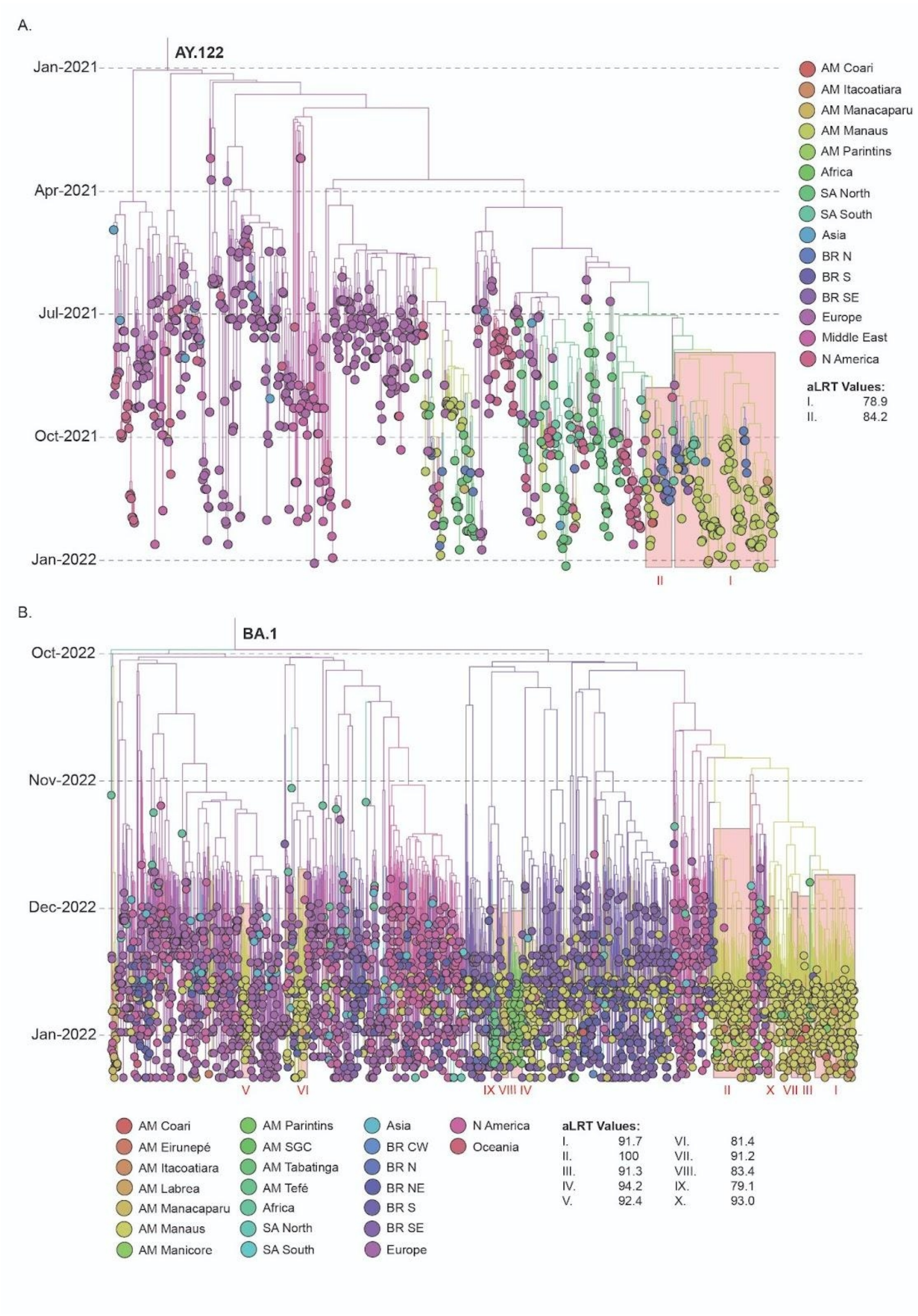


**Figure S3: Spatio Temporal Characterization of lineages AY.122 and BA.1 in Amazonas State.** Time-scaled MCC tree of SARS-Cov-2 lineages AY.122 ($n_{TOTAL}$ = 839) and BA.1 ($n_{TOTAL}$= 3,445). Branches are colored according to their most probable location, based on the color scheme shown in the legends. Large clusters in each lineage are indicated alongside the aLRT support obtained in the previous ML analysis. AM: Amazonas, N: north, NE: northeast, S: south, SA: South America, SE: southeast, CW: central-west.

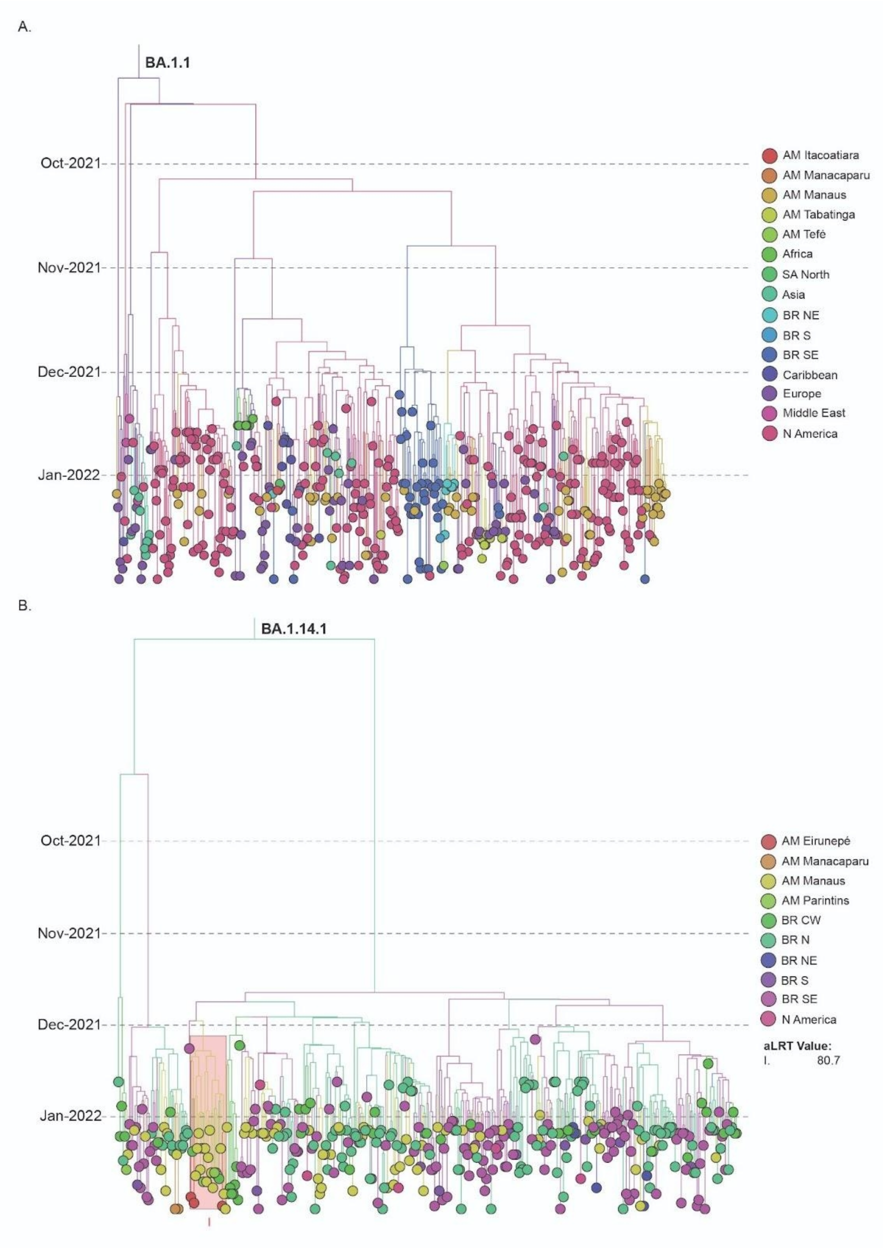


**Figure S4: Spatio Temporal Characterization of lineages BA.1.1 and BA.1.14.1 in Amazonas State.** Time-scaled MCC tree of SARS-Cov-2 lineages BA.1.1 ($n_{TOTAL}$ = 478) and BA.1.14.1 ($n_{TOTAL}$= 394). Branches are colored according to their most probable location, based on the color scheme shown in the legends. Large clusters in each lineage are indicated alongside the aLRT support obtained in the previous ML analysis. AM: Amazonas, N: north, NE: northeast, S: south, SA: South America, SE: southeast, CW: central-west.

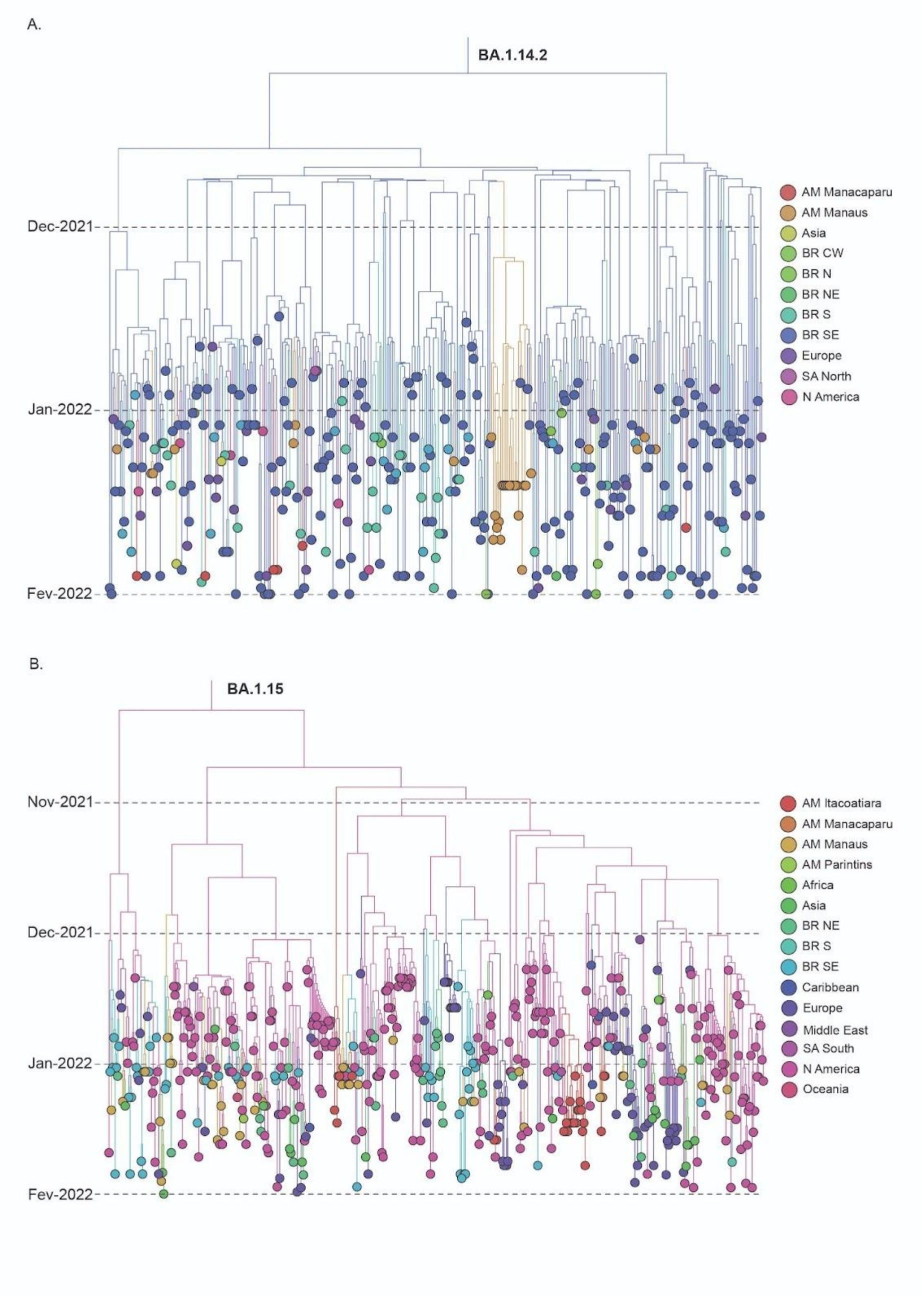


**Figure S5: Spatio Temporal Characterization of lineages BA.1.14.2 and BA.1.15 in Amazonas State.** Time-scaled MCC tree of SARS-Cov-2 lineages BA.1.14.2 ($n_{TOTAL}$ = 363) and BA.1.15 ($n_{TOTAL}$= 526). Branches are colored according to their most probable location, based on the color scheme shown in the legends. AM: Amazonas, N: north, NE: northeast, S: south, SA: South America, SE: southeast, CW: central-west.

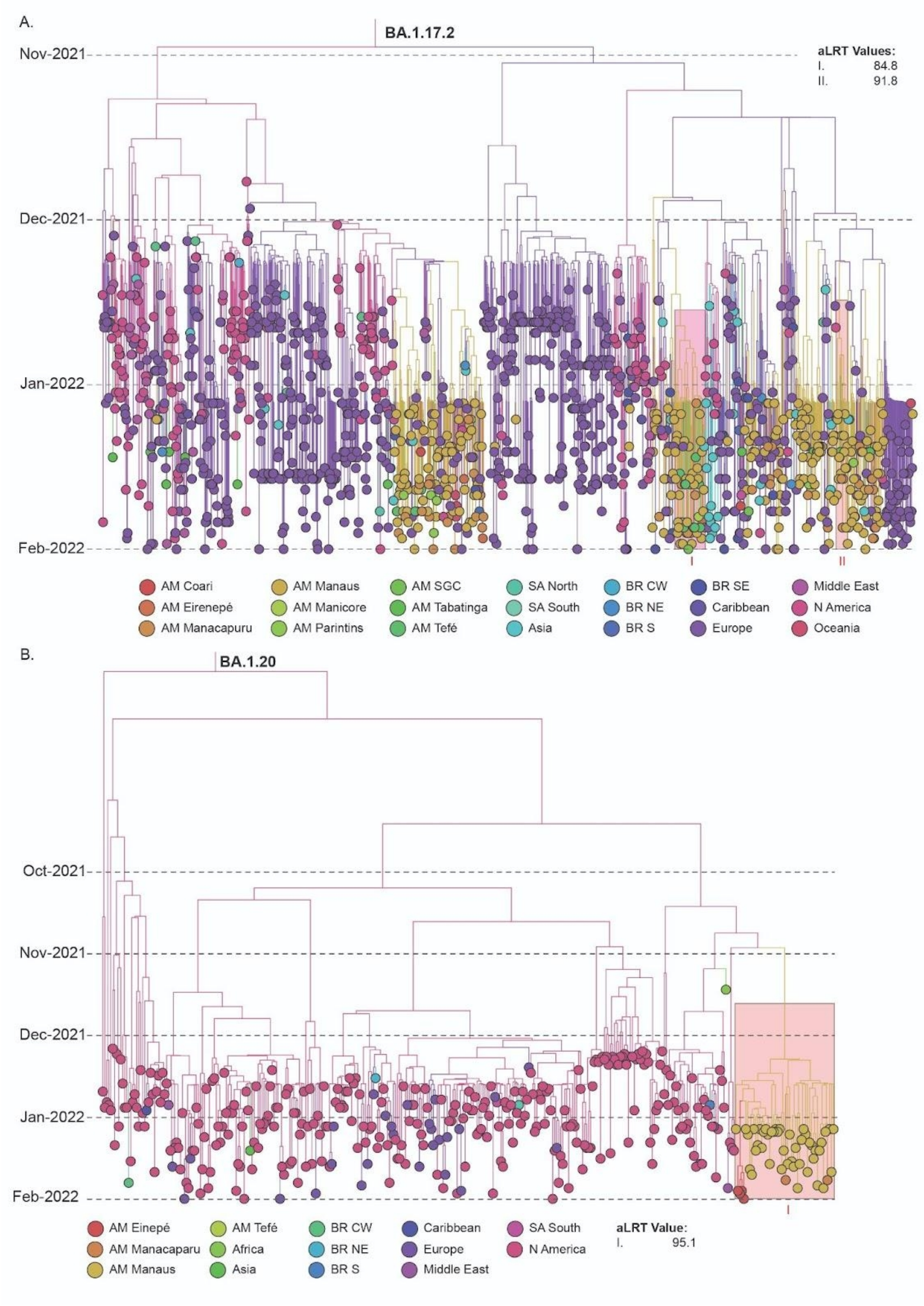


**Figure S6: Spatio Temporal Characterization of lineages BA.1.1 and BA.1.14.1 in Amazonas State.** Time-scaled MCC tree of SARS-Cov-2 lineages BA.1.1 ($n_{TOTAL}$ = 478) and BA.1.14.1 ($n_{TOTAL}$= 394). Branches are colored according to their most probable location, based on the color scheme shown in the legends. Large clusters in each lineage are indicated alongside the aLRT support obtained in the previous ML analysis. AM: Amazonas, N: north, NE: northeast, S: south, SA: South America, SE: southeast, CW: central-west.

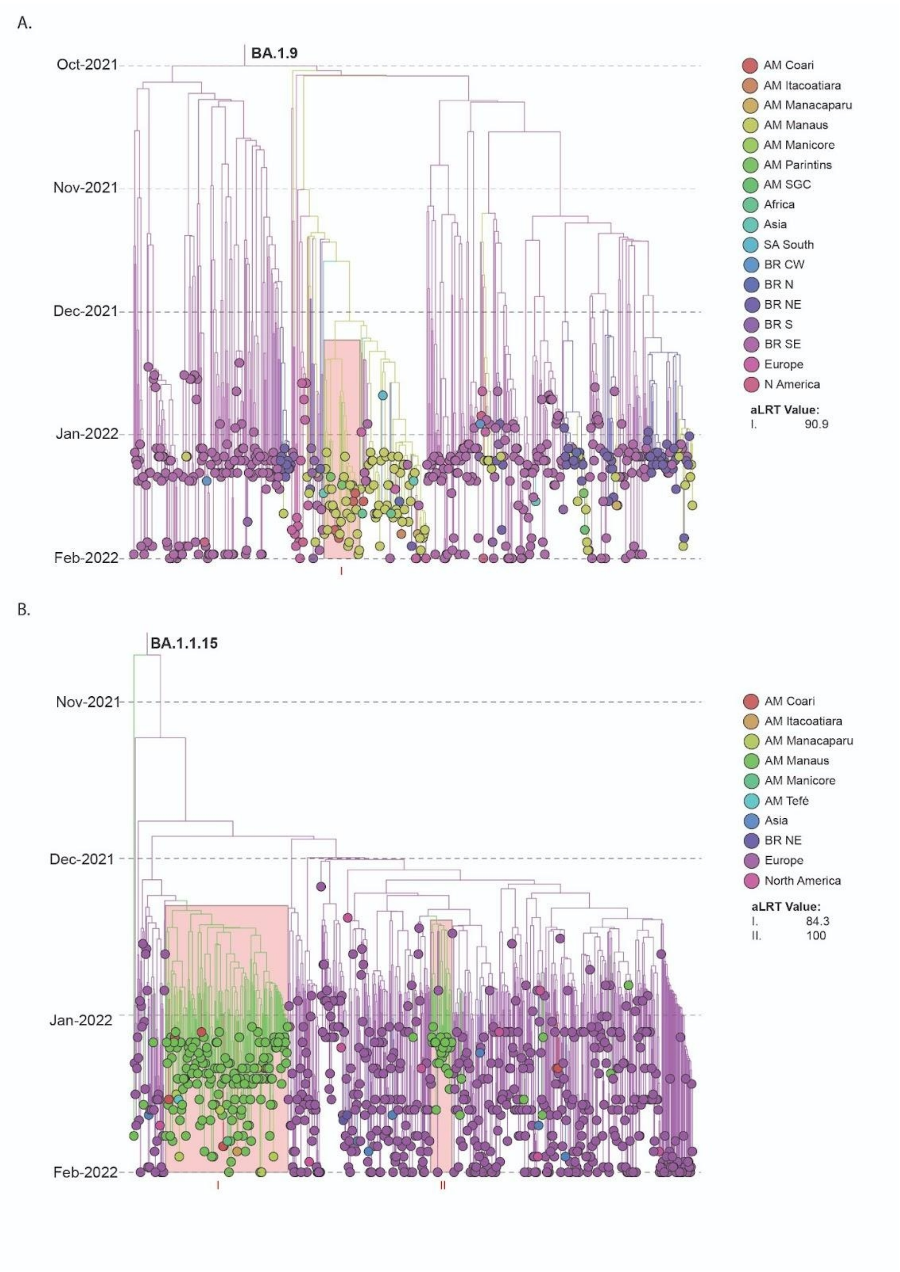


**Figure S7: Spatio Temporal Characterization of lineages BA.1.9 and BA.1.1.15 in Amazonas State.** Time-scaled MCC tree of SARS-Cov-2 lineages BA.1.1 ($n_{TOTAL}$ = 478) and BA.1.14.1 ($n_{TOTAL}$= 394). Branches are colored according to their most probable location, based on the color scheme shown in the legends. Large clusters in each lineage are indicated alongside the aLRT support obtained in the previous ML analysis. AM: Amazonas, N: north, NE: northeast, S: south, SA: South America, SE: southeast, CW: central-west.
